# Program Cost and Return on Investment of a Remote Patient Monitoring Program for Hypertension Management

**DOI:** 10.1101/2025.01.29.25321334

**Authors:** Donglan Stacy Zhang, Laure Millet, Brandon K. Bellows, Sarah Lee, Devin Mann

## Abstract

**Objective:** To evaluate the program costs and financial sustainability of a remote patient monitoring for hypertension (RPM-HTN) program implemented in the cardiology practice of a large healthcare system.

**Study Design:** This economic evaluation utilized field observation, interviews, literature review, and quantitative analysis to assess RPM-HTN from March to June 2024 at New York University Langone Health

**Methods:** A costing tool was developed to quantify program costs, including personnel, start-up, equipment, and supply expenses, expressed in 2024 USD. Reimbursement rates were estimated using the 2024 Medicare Physician Fee Schedule. The return on investment (ROI) was calculated as the ratio of net return to program costs. Univariate sensitivity analyses evaluated the impact of varying a single parameter at a time on ROI.

**Results:** The average cost of RPM-HTN was $330 per patient (range: $208–$452), with an annual program cost of $33,000 (range: $20,785–$45,168) for 100 patients enrolled from the Cardiology Division. Key expenses included data review by nurse practitioners ($172/patient), blood pressure device costs ($48/patient), and nurse-patient communication ($36/patient). ROI averaged 22.2% at 55% patient compliance with the RPM-HTN program. This ROI ranged from-11.1% (assuming program costs of $452) to 93.3% (assuming program costs of $208) per patient. ROI was most sensitive to changes in data review costs, insurance reimbursement, patient compliance, and device setup.

**Conclusions:** The RPM-HTN program demonstrated positive ROI, indicating financial sustainability in a large urban healthcare system. Improving patient compliance with the program and reducing human resource costs are critical for scaling RPM-HTN programs effectively.

## INTRODUCTION

Hypertension, defined as systolic blood pressure (BP) ≥130 mm Hg or diastolic BP ≥80 mm Hg before treatment, affects 50.4% of males and 43.0% of females aged 20 years or older in the United States.^1,2^ Hypertension is a major cause of cardiovascular disease and the leading cause of death in the U.S., leading to immense health and economic burden.^1,3^ Typical management of hypertension occurs during office visits with primary care providers where BP is measured and antihypertensive medication changes may be made. Remote patient monitoring for hypertension (RPM-HTN) combines measurement of BP at home with electronic transmission of BP readings to providers and telehealth services.^4–6^ Several randomized controlled trials have demonstrated that RPM-HTN can be effective and cost-effective in managing hypertension, enhancing BP control, and mitigating cardiovascular disease events.^7–12^

Owing to the significant shifts in telehealth brought about by the COVID-19 pandemic, RPM- HTN is increasingly becoming an integral part of routine healthcare.^13^ Since 2017, NYU Langone Health (NYULH) has implemented an RPM-HTN program across diverse clinical settings. The growing ambulatory RPM program has enrolled over 11,000 patients in home-based monitoring of at least one physiological parameter, with over half including a HTN component.^14^ Unlike other health systems that focus on centralized RPM programs for specific high-need populations, NYULH emphasizes building tools and workflows for practices interested in using RPM. This unique approach supports diverse clinical departments in integrating RPM into their patient care strategies.

Scaling up RPM-HTN requires estimates of the cost to implement and run a program, including the resources needed to integrate RPM-HTN into routine healthcare infrastructure and evolving workflows.^15,16^ In addition, regulatory considerations, particularly payment and reimbursement policies, significantly impact the adoption and potential short-term return on investment (ROI) needed to ensure financial sustainability of RPM-HTN. Not all private insurance or Medicaid programs reimburse RPM services. The Centers for Medicare and

Medicaid Services (CMS) reimburse RPM-HTN usage, but billing for RPM is complex, leaving certain services unreimbursed, which may lead to a negative ROI and deter systems from adopting these technologies.^17^ Understanding the drivers of program costs and ROI is essential for payers to establish or modify payment rates and adopt value-based models that align financial incentives with value-based care initiatives.

To address these evidence gaps, we investigated the program costs and ROI of an RPM-HTN program from the perspective of the Cardiology Department at NYULH.

## METHODS

To estimate the costs associated with implementing and operating RPM-HTN, we examined the workflow of the program and used an activity-based micro-costing approach, which identifies and values every resource utilized. We classified costs into several categories: personnel, program startup, equipment, supply, and other miscellaneous costs required for the implementation and operation of RPM-HTN, modified from a standardized costing tool.^18^ We collected cost data through an on-site visit (observing in the Cardiology Division, March-May 2024), staff reporting derived from a survey (**Appendix Table 1**). ROI analysis inputs were sourced from observations, provider self-reports, NYULH EHR records, and reliable local and national websites for salary and payment rates for RPM services (**Table 1**).

**Table 1.** ROI Analysis: Data Inputs and Sources.

| Inputs | Sources |
| --- | --- |
| Personnel cost | On-site visit, staff self-reports, EHR records, and average salary for each provider type in New York State (e.g., hourly rate) |
| Start-up cost | On-site visit, staff self-reports |
| Equipment cost | Receipts, staff self-reports |
| Supply cost | Receipts, staff self-reports |
| Patient characteristics and factors (e.g., duration, emergency, difficulty using devices, type of payment) | EHR records, staff self-reports |
| Patient compliance rate (e.g., submitting BP readings) | EHR records, staff self-reports |
| Payment rate | CMS physician fee schedule |

### RPM-HTN Workflow

The NYULH Cardiology Division enrolled 100 new patients whose BP was not controlled into the RPM-HTN program. To analyze program workflow and activities, the staff in the division provided initial data during an on-site visit. Any remaining questions were then addressed through emails and virtual meetings. The RPM-HTN program workflow included five steps: Patient enrollment, Device setup, Data monitoring, Follow-up, and Discharge from the RPM program (**Figure 1**). The cardiologist was responsible for enrolling patients, after which the nurse practitioner (NP) and medical assistant (MA, who also serves as the research coordinator) assisted patients in signing the consent forms, setting up the devices, and providing educational materials on how to use the BP devices and sync data with the Epic-enabled smartphone application. After the patient began using the device to monitor their blood pressure, the NP checked the data weekly to ensure it was being submitted regularly. The NP may also call the patient to take or adjust medications as needed, while the MA assists with troubleshooting any issues the patient encounters with the device or app. In the rare event of an emergency situation, the NP would call the patient to schedule an appointment with the cardiologist. Once a patient’s BP has stabilized for at least one month, they will be discharged from the program.

**Figure 1.**
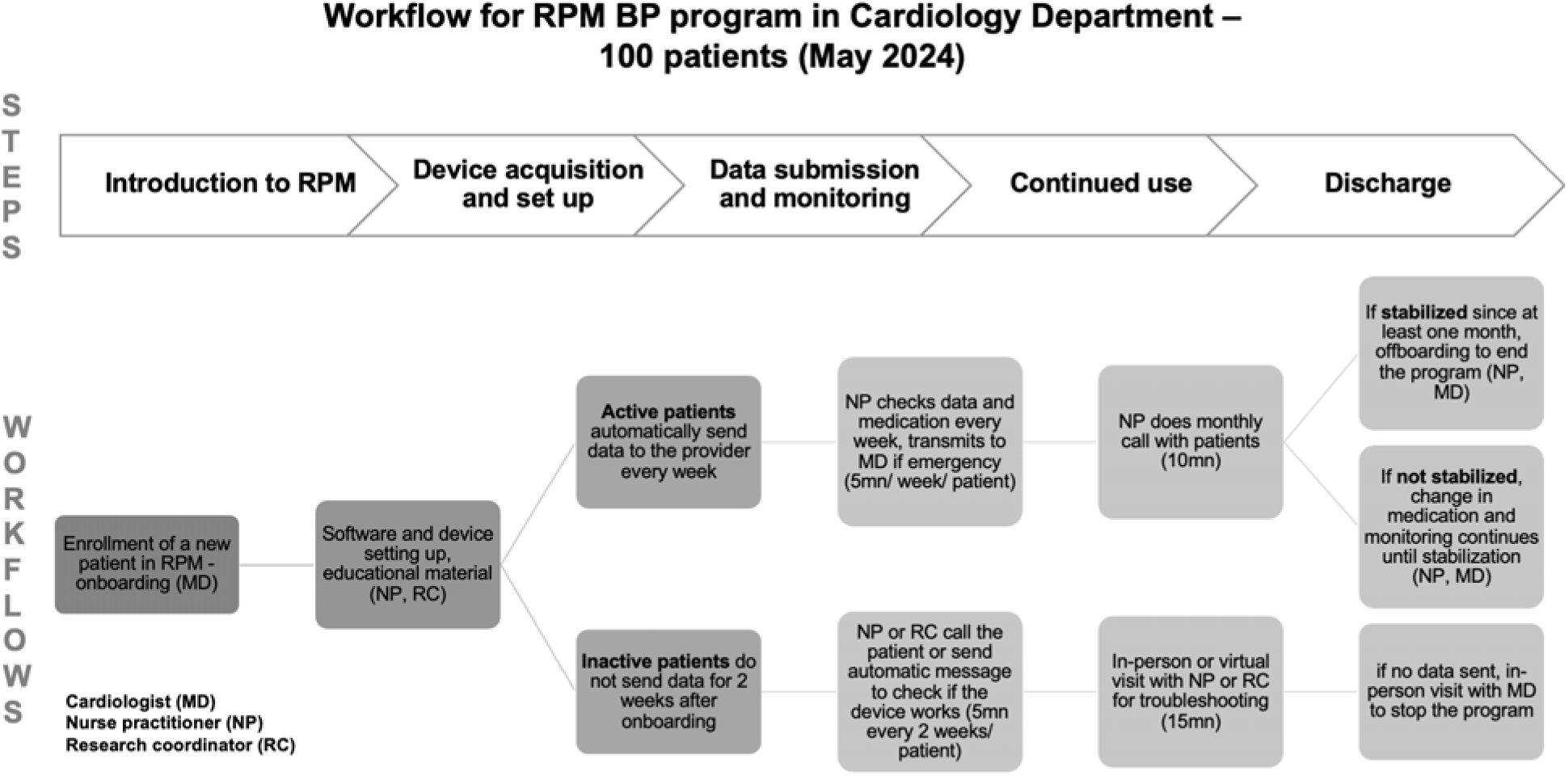
Remote Patient Monitoring Workflows

### Valuation of Personnel Costs

We used a bottom-up approach to estimate staff time, primarily relying on survey responses to determine the time spent on each activity, the number of patient encounters, and the number of patients per month. Time was reported as the average duration for each patient encounter, along with the minimum and maximum values.

The data were subsequently presented at the hospital’s monthly RPM strategy meeting for discussion and to address any additional questions. We followed up with a virtual meeting with the staff to verify the collected data. Personnel costs were estimated based on average salaries and fringe benefits for each provider type in the New York metropolitan area, including cardiologists, nurse practitioners, nurse assistants, and administrative staff. The hourly rates for each provider type were obtained from Indeed, a job search platform website that provides salary information.^19^ Reported staff time was then added and multiplied by these hourly rates.

All costs were adjusted to 2024 US dollars using the Personal Health Care Expenditure component of the National Health Expenditure Accounts, following the guidelines recommended by the Agency for Healthcare Research and Quality.^20^

### Program Startup Costs

An RPM program’s startup costs may include equipment, infrastructure, and personnel expenses. We classified startup items such as BP monitors and software as equipment costs, while information technology (IT) infrastructure costs, such as Epic system upgrades, were centralized and did not incur expenses for the Cardiology Division. Consequently, the preliminary startup costs for the program primarily consisted of personnel-related expenses, including staff training. Since no new staff were hired to manage hypertension patients using RPM-HTN, the startup costs were mainly associated with training. We calculated the number and duration of training sessions for the care team and annualized these costs.

### Equipment Costs

The hospital purchased equipment, including the EHR-embedded application Validic, a middleware to connect home BP devices to the NYU EHR. The BP devices were purchased by the department and distributed to patients upon enrollment. These devices were recommended to be returned upon patient discharge and then reused for newly enrolled patients. We obtained the purchase costs for Validic and the BP devices (at discounted prices) from the Finance Department’s reports and validated these prices with receipts. Since the Validic equipment would ideally be used for multiple years, we annuitized the costs. The useful life of the equipment was determined using guidance from the Internal Revenue Service (IRS), and it was determined that the EHR-embedded app had a useful life of 10 years.^21^ We used the straight-line method to depreciate the equipment over its useful life.^22^ The value of the equipment over its useful life was then divided by an annuity factor, using a 3% discount rate, and further divided by the estimated number of patients in the health system who used the Validic app.

### Supply Costs

Supplies and materials included educational materials for patients on how to set up the devices, sync data, and monitor their BP, translation materials for patients who do not speak English or prefer Spanish as their primary language, and written consent forms for enrolling patients into the RPM-HTN program. Since these costs were incurred only once during patient enrollment, we estimated them on a per-patient basis.

### ROI Analysis

Revenue to offset program costs comes from insurance reimbursements and patient out-of-pocket payments. Patients were in the program for an average duration of four months.

Payment rates set by private payers varied substantially, while Medicare established a physician fee schedule, allowing providers to use CPT codes to bill for RPM services.^23^ These codes include: CPT 99453, covering initial setup and patient education on RPM equipment ($19). CPT 99454, used for supplying devices, is billable once per patient per month with a minimum of 16 BP recordings ($50). As a result, patient compliance in submitting their BP records directly affects the revenue generated from RPM-HTN services. CPT 99091, for collecting and interpreting patient data for a minimum of 30 minutes per month ($54). CPT 99457, covering the first 20 minutes of clinical staff communication with the patient each month ($49). CPT 99458, for each additional 20 minutes of interactive communication beyond the initial 20 minutes covered by CPT 99457 ($40). For Medicare patients receiving services at a federally qualified health center, CPT G0511 can be billed under chronic condition management with RPM monthly services ($73). Moreover, the CMS physician fee schedule rates vary by state. In New York, adjustments for labor costs and practice expenses use a rate of 1.061, while adjustments for non-labor costs use a rate of 1.184. Therefore, we adjusted the payment rates in the ROI analysis using a range from 1.061 to 1.184. ^23^

We used the Medicare payment rate to estimate the total revenue generated from the RPM program per patient and for the department per year. We then tested different reimbursement structures in a sensitivity analysis.

The annual ROI was estimated using the following formula:^24^

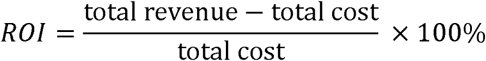

The numerator represents the difference between total revenue and total cost, or the expected value (EV) of the program, while the denominator is the program’s total cost. This short-term ROI analysis did not account for longer-term health benefits (e.g., improved medication adherence, higher control rates, prevention of cardiovascular events), opportunity costs (e.g., investments in other potentially beneficial programs), or broader health and societal gains (e.g., savings from reduced travel time for care) associated with RPM-HTN.

### Sensitivity Analysis

We assessed the robustness of the ROI analysis for RPM-HTN through a series of univariate sensitivity analyses. Specifically, we accounted for uncertainties related to personnel costs (data viewing, device setup and follow-up costs), including the estimated average time spent with patients per encounter and the hourly rates for the cardiologist, NP and MA, patients’ compliance rate in submitting BP readings, and insurance reimbursement rates, which included the lowest payment rate using CPT code G0511, higher payment rates used by private payers than Medicare, and patient self-payment. Additionally, we analyzed how uncertainties in clinical factors, such as the percentage of patients experiencing emergency situations, affected the EV of the ROI estimate for RPM-HTN.

## RESULTS

Overall, the average cost per patient enrolled in the RPM-HTN program was estimated at $330 (range: $208–$452). The projected total cost for the NYULH Cardiology Division to manage 100 patients was $33,000 (range: $20,785–$45,168) (**Table 2**). Reviewing patient home BP data by the NP was projected to be the most costly component, with an average per-patient cost of $172 (range: $86–$258). The cost of BP devices ($48) as well as monthly communication with patients by the NP ($36; range: $24–$48) were estimated to be the next two highest program costs. The cost to integrate BP monitoring data with the Epic system using the software was estimated to be $23. Other costs, such as start-up and supply costs, were generally low. Start-up costs included staff training to implement the program, which lasted 30 minutes, with a per-patient cost of $0.42, and supply costs related to printed educational materials and translation services were estimated at $1.70 per patient.

**Table 2.** Cost of the Remote Patient Monitoring Programs for Hypertension Management in A Cardiology Department (Number of Enrollment = 100)

| | | Cost Per Patient (\$) <sup>d</sup> | | | Average Department Cost (\$) <sup>d</sup> | | |
| --- | --- | --- | --- | --- | --- | --- | --- |
|  | Provider <sup>a</sup> | Average | Minimum | Maximum | Average | Minimum | Maximum |
| Personnel cost |  |  |  |  |  |  |  |
| Patient enrollment | MD | 29 | 15 | 44 | 2917 | 1458 | 4375 |
| Device setup | NP | 11 | 5 | 16 | 1,073 | 537 | 1610 |
| Data reviewing | NP | 172 | 86 | 258 | 17,171 | 8585 | 25756 |
| Communication <sup>b</sup> | NP | 36 | 24 | 48 | 3564 | 2376 | 4752 |
| Troubleshooting | NA | 1 | 1 | 2 | 128 | 102 | 154 |
| Administration <sup>c</sup> | NP | 8 | 4 | 12 | 794 | 397 | 1191 |
| Start-up cost |  |  |  |  |  |  |  |
| Staff training | NP, NA | 0.42 | N/A | N/A | 42 | N/A | N/A |
| Equipment cost |  |  |  |  |  |  |  |
| Software/vendor | N/A | 23 | N/A | N/A | 2,325 | N/A | N/A |
| BP Devices | N/A | 48 | N/A | N/A | 4793 | N/A | N/A |
| Supply cost |  |  |  |  |  |  |  |
| Written consent forms | N/A | 0.3 | N/A | N/A | 30 | N/A | N/A |
| Educational materials | N/A | 0.7 | N/A | N/A | 70 | N/A | N/A |
| Translational services | N/A | 0.7 | N/A | N/A | 70 | N/A | N/A |
| **Total cost** | N/A | 330 | 208 | 452 | 32977 | 20785 | 45168 |

Note: a. Providers involved in the RPM-HTN program include a cardiologist, a nurse practitioner and a certified nursing assistant.
b. Communication refers to nurse-patient interactions about whether BP is normal, if medication adjustments are needed, and/or if an in-person visit is necessary.
c. Administrative activities include signing off patient consent forms and billing for services.
d. Cost estimates rounded to an integer.

The ROI results are shown in **Table 3**. Current patient compliance in submitting BP readings was estimated at 55%. When applied to both cost and reimbursement estimates (e.g., patients noncompliance makes the service non-billable), the average ROI is 22.2%, meaning that for every dollar invested in the RPM-HTN program, there is a return of $1.22. This ROI ranges from-11.1% (assuming program costs of $452) to 93.3% (assuming program costs of $208) per patient. If we estimated the ROI at the division level, the ROI could be 19.5% in the base case scenario, with a range from-12.8% to 89.5%. Lower program costs resulted in more favorable ROI ratios, indicating better financial performance and value generation from the RPM-HTN program.

**Table 3.** Return-On-Investment Analysis of the Remote Patient Monitoring for Hypertension Management.

| **Cost** | **Compliance rate <sup>a</sup>** | **Medicare Insurance Reimbursement** | **Expected value** | **Return on investment ratios** |
| --- | --- | --- | --- | --- |
| Per patient (average duration: 4 months) |  |  |  |  |
| Average: \$329 | 55% | \$402 | \$73 | 22.2% |
| Minimum: \$208 | 55% | \$402 | \$194 | 93.3% |
| Maximum: \$452 | 55% | \$402 | -\$50 | -11.1% |
| Division (100 patients; average duration: 4 months) |  |  |  |  |
| Average: \$32,977 | 55% | \$39,396 | \$6419 | 19.5% |
| Minimum: \$20,785 | 55% | \$39,396 | \$18,611 | 89.5% |
| Maximum: \$45,168 | 55% | \$39,396 | -\$5772 | -12.8% |

Note: The compliance rate is estimated to be around 55%.

Sensitivity analyses of key parameters in the ROI estimate numerator (expected value, EV) are presented in **Figure 2** as a tornado diagram. The diagram illustrates changes in EV when each variable is adjusted to its maximum and minimum values, emphasizing their relative importance in the ROI estimate. The most influential factor was the cost of data viewing, ranging from $86 to $258 based on the NP’s weekly BP review time and hourly rate, leading to an EV range of-$13 to $159 (**Appendix Table 2**). The second most influential factor was program payments, varying from $292 (FQHC rate) to $525 (higher-than-Medicare rate), resulting in an EV range of-$16 to $77 per patient. The third key factor was patient compliance, ranging from 0.4 to 0.8, with an EV range of $73 to $128. Device setup costs, varying from $93 to $133, influenced the EV from $53 to $93. In addition, the proportion of patients who initially failed to submit data due to technical issues but became compliant after troubleshooting increased the EV. Similarly, the cost of NP follow-ups, ranging from $50 to $100, resulted in an EV range of $54 to $87. The probability of emergency events also affected the EV but had a smaller impact compared to other factors.

**Figure 2.**
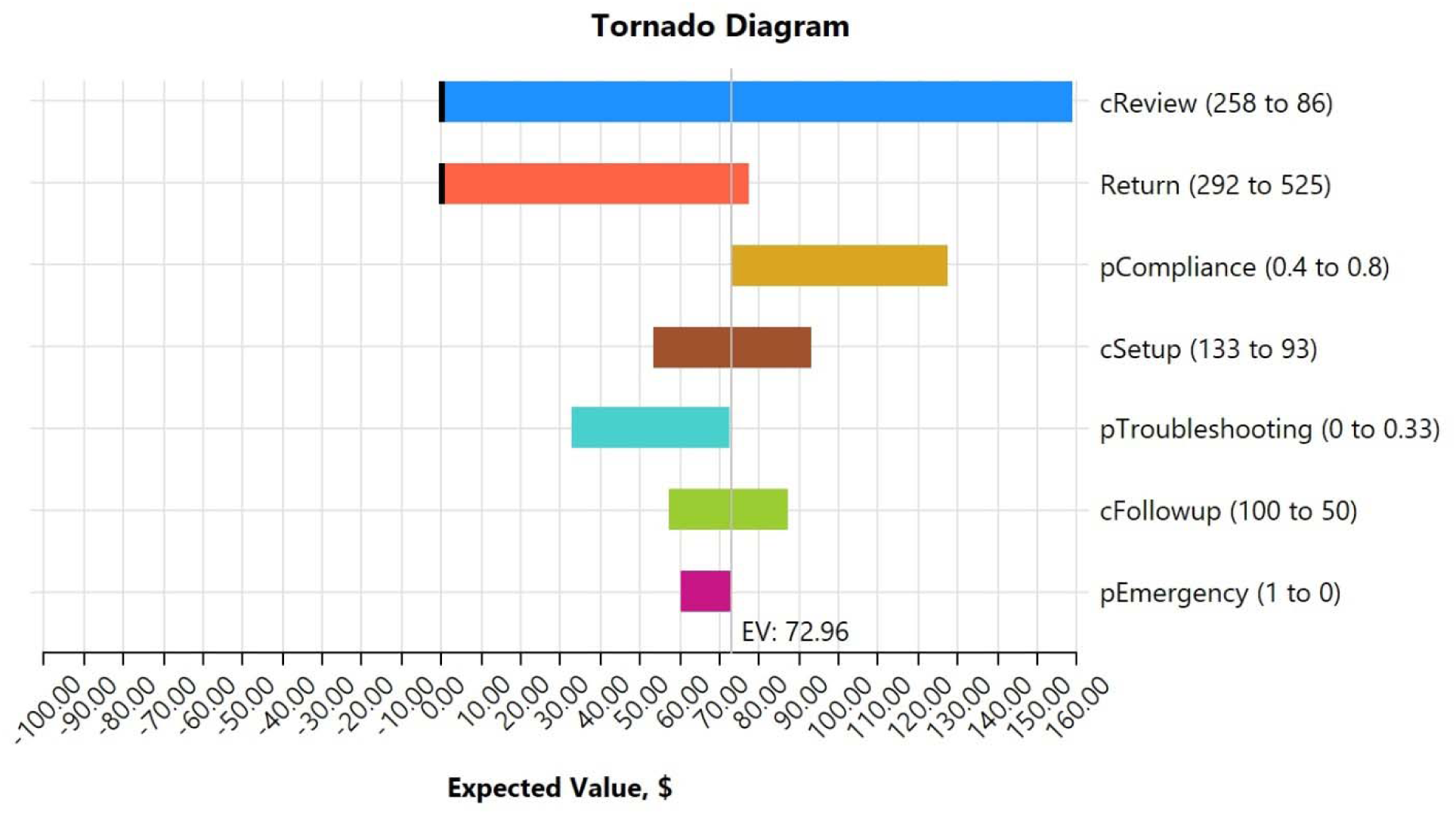
Sensitivity Analysis of Key Parameters in the Estimated EV (Tornado Diagram)

## DISCUSSION

In this study, we estimated the program cost of RPM-HTN in the Cardiology Division at NYU Langone Health and found that, on average, the program cost was $330 per person. The estimated ROI was positive at 22.2%, even with an average compliance rate of around 55%, indicating that the program is generating a profit above the costs, making it a financially sustainable program.

The Community Preventive Services Task Force recommends RPM to reduce BP, citing its effectiveness and cost-effectiveness.^25^ However, the lack of guidelines for RPM-HTN workflows limits its widespread adoption due to concerns about cost and sustainability. Our findings, based on an RPM program in a large healthcare system, show that RPM-HTN can be a key component of healthcare delivery, leading to cost savings and enhance efficiency.

Organizational workflows significantly influence the ROI of RPM, with integration impacting costs for resources, software, and training.^7,25,26^

Regarding costs, we found that more than half of the RPM-HTN program’s expenses ($171.71 per patient, 52.07%) were due to data-viewing tasks overseen by the NP. This reflects the significant time and effort needed to monitor and analyze patient data for hypertension management. Data review was the most time-consuming task, ensuring BP records were submitted regularly, accurate, and monitored for control.^26^ This process is vital for successful home-based monitoring, medication administration, detecting masked hypertension, and avoiding higher-acuity care.^25–28^ Research found that intervention protocols triggering patient-provider interactions on an as-needed basis were associated with lower costs and greater effectiveness.^25^

Our study found that although staff training was the most expensive start-up cost, the cost per patient remained low. In addition, startup costs such as purchasing EHR-embedded software (Validic), Bluetooth-enabled BP monitors, and training can be minimized when scaling up the RPM-HTN program to other clinical divisions.^14^ A loaner program for BP monitors could reduce costs by recycling devices from discharged patients to new patients.^29^ Moreover, some patients without BP cuffs or insurance coverage could benefit from a loaner program, further reducing financial barriers for patients.^29^ As the program expands, costs will rise, making reuse and loaner programs important. Additionally, automation and delegation strategies may reduce patient follow-up costs. For example, the healthcare system is exploring artificial intelligence to handle some tasks currently performed by NPs,^30^ such as sending personalized reminders to measure BP or take medications, and assisting with scheduling virtual appointments for elevated BP, which may improve patient medication adherence and compliance in remote monitoring.^31^

Patient compliance is critical for the ROI of the RPM-HTN program.^25^ Non-compliance leads to lost revenue, and further research is needed to understand patient engagement behaviors.

One way we propose to enhance patient experience and engagement is by incorporating simplified user interfaces, improve patients’ onboarding experiences, and providing multilingual resources.^32,33^ Studies show that personalized care strategies, such as tailored feedback, reminders, and incentives, can significantly boost patient engagement.^31,34,35^ Data analytics can track usage patterns and engagement, aiding ROI analysis and interventions to boost compliance.

Economic evaluation is important to set reimbursement policies. We calculated ROI using CMS billing codes and Medicare reimbursement rates, which, while providing stability, impose restrictive quotas such as requiring 20 minutes of monthly patient interaction and prohibiting simultaneous billing for Remote Physiologic and Therapeutic Monitoring. The variety of insurance types among Cardiology Division patients, including private insurance and Medicaid, may not provide the same reimbursement as Medicare. Rigid reimbursement rules, such as requiring ≥16 data uploads per month or interactive encounters, limit RPM growth at scale.

Value-based payment models, such as shared savings programs or capitated insurance, could incentivize high-quality, cost-effective care, particularly for patients with multiple chronic conditions.^36,37^

This study has several limitations. First, the cost and ROI estimates reflect only one cardiology division and may not be generalizable to various patient populations, such as pregnant women with hypertensive disorders. Second, cost estimates are from the healthcare system’s perspective; future studies should integrate cost estimates from the patient’s and payer’s perspectives. Third, we used Medicare reimbursement rates to estimate the return, but many patients have private insurance or Medicaid, which have varied reimbursement policies. Future research should examine the impact of various payment models on RPM-HTN adoption using claims databases. Finally, we assessed short-term cost-effectiveness, as patients were in the program for an average of four months. Long-term evaluations should be incorporated as the program continues.

In conclusion, our study examined the operational workflow, resource allocation, and ROI of an RPM-HTN program in a large healthcare system. We found the program generated positive ROI and identified factors for improving efficiency. Future research should explore the barriers to scaling the program to other clinical departments and patient populations and investigate the long-term financial suitability of RPM-HTN and its alignment with reimbursement policies to improve hypertension care.

## Summary of the article

This study assesses the costs and financial sustainability of a Remote Patient Monitoring program in cardiology, revealing a positive return-on-investment and identifying key cost drivers.

## Take-away Points

- The Remote Patient Monitoring program achieved a positive return on investment (22.2%), indicating its potential for cost-effective hypertension management in large healthcare systems.
- Key cost drivers include data review by nurse practitioners and equipment expenses. Strategies like task automation and device recycling can potentially improve financial performance.
- Increasing patient adherence to submitting blood pressure readings is essential to maximize reimbursement and return on investment.
- Modifying insurance reimbursement policies to reduce administrative complexity and incentivize value-based care models has the potential to promote Remote Patient Monitoring adoption and scalability.

## Data Availability

All data produced in the present study are available upon reasonable request to the authors

## Acknowledge

The authors would like to express our sincere gratitude to Trisha Aloquina, Ayanna Horsford, and the other staff at NYU Langone Health who assisted us in collecting data for the cost analysis of the remote patient monitoring program. We also acknowledge the following funding support: National Science Foundation Grant No. 2129076 (Mann) and National Institutes of Health Grant No. R01MD013886 (Zhang).

